# Presumptive Multidrug-Resistant *Escherichia coli* Isolated in Drinking Water and Soil Sources from Kadamaian, Sabah

**DOI:** 10.1101/2022.03.20.22272634

**Authors:** Noor Farhani Atiqah Jamrin, Norshafiqa Suhaimi, Muhammad Zul Izlan Zulkifli, Nur Athirah Yusof

## Abstract

Multi-drug-resistant *Escherichia coli* poses a great threat to human health. *E. coli* is often used as fecal indicator bacteria to study the sources and fate of fecal contamination in the environment. This study focused on water and soil samples in tourist parks around Kadamaian, Kota Belud from three different villages of Kampung Napungguk, Kampung Pinalabuh and Kampung Tinata. From 23 of tested presumptive *E. coli* isolates, the percentage of antimicrobial-resistant against a series of antibiotic concentrations ranging from 25, 50, 100 to 500 μg/ml showed resistance towards Chloramphenicol (91.3%), Penicillin (82.6%), Tetracycline (73.9%) and Kanamycin (30.43%). This finding suggested the presence of multi-drug resistance (MDR) of presumptive *E. coli* from soil samples of Kampung Napungguk and Kampung Tinata. This study alarms the presence of MDR which need further research on identifying the bacterial strains using molecular methods. The findings can then be used to create awareness of potential disease outbreaks in the tourist areas.

## Introduction

According to the World Health Organization, unsafe water has caused 80% of all diseases around the world (Ellis & Schoenberger, 2017). About 10% of the diseases were related to water diseases such as diarrhea which has become a plaque and burden the developing countries particularly (Mari et al., 2018). According to Odonkor & Addo (2018) for many decades, *E. coli* has been the top-tier indicator of faecal contamination in the water quality. Basically, during rainfalls, these coliforms are washed away into the stream, river, lake and groundwater. Thus, untreated drinking water coming from these sources have a high possibility of containing many coliforms including *E. coli* (Odonkor & Addo, 2018). According to Arsene-Ploetze et al., 2018; Katakweba et al., (2018), *E. coli* exhibit a significant reservoir of genes that code for antimicrobial drug resistance and therefore, it can be a useful indicator for resistance in the communities of bacteria. These antibiotic-resistance genes are conferring to a variety of antibiotics that have been found in a wide range of water environments especially drinking water in both developed and developing countries (Marathe et al., 2017; Mezrioui & Baleux, 1994). Furthermore, this becomes the main risk to public health as these resistance genes are shifted from environmental bacteria to human pathogens. According to Bengtsson-Palme & D. J. Larsson, 2016; Pan & Chu, (2018), it has been documented that drinking water has become the medium for the transportation of microbial pathogens that cause several illnesses.

The study conducted was aimed to isolate bacteria that were presumptive to be *E. coli* that contaminated local recreational areas around Kadamaian, Kota Belud, Sabah using the selective culture media method. The objective of the research was to identify the occurrence of antibiotic resistance of bacteria that were presumed to be *E. coli* that could be found in the recreational areas. Although there have been many studies conducted on the occurrence of *E. coli* in food products, this research is the first research that attempts to study the microbial communities around the local recreational areas in Kadamaian, Kota Belud, Sabah. According to (Geraldine, 2019), Kadamaian is the fast-growing rural tourism spot in Sabah which has received over 100,000 tourists and visitors every year. This beautiful place is located in Kota Belud, which offers a variety of rural eco-tourism products such as waterfalls, pristine rivers, forests and also hills.

Due to these attractions, which has attracted many domestic and foreign tourists from many places, Kadamaian water sources have a high possibility to be contaminated by *E. coli* which are commonly found in the lower intestine of humans and animals (Jang et al., 2017). Besides, the unsystematic sanitation among the rural community also could lead to the outbreak of pathogenic strains of bacteria, especially among the low immunity categories such as children and the elderly. Data from this study could provide the foundation for further research in which can be utilized by the public health bodies and other governmental organisations to raise awareness among the local communities so that prevention measures can be taken on the type of bacteria strains and their antibiotic-resistance attributes.

## Materials and methods

### Sample collection sites

In this study, the sites chosen were recreational areas located at the tourists’ spots around Kadamaian, Kota Belud, Sabah. Selected sites were Kampung Tinata (T), Kampung Napungguk (N) and Kampung Pinalabuh (P) in which located in Kadamaian, Kota Belud. All samples including water and soil samples were taken from the locations that were representative of the water sources or distribution networks from which water was regularly used by the inhabitants. Before the sampling activity started, observations on the sanitary conditions of that area were done to eliminate any possible sources of contamination that could affect the quality of the samples. A total of 19 samples consisting of water and soil samples were collected from these three areas. All water sampling and preservation were performed according to the standard methods for the examination of water and wastewater (Burgmann et al., 2018, Jang et al., 2017).

### Isolation of presumptive *E. coli* from water samples

For the isolation of presumptive *E. coli* from the water samples, 100 μl of a water sample from each site was spread evenly on the Eosin methylene blue (EMB) media. For the isolation of presumptive *E. coli* from the soil samples, 1 g of each soil sample was dissolved in 10 mL of sterile peptone broth. Samples were vortexed vigorously for a minute. Next, several dilutions were performed: 10^2^, 10^3^, 10^4^, 10^5^, 10^6^, 10^7^ and 10^8^. Dilution was done by adding 1 mL of water samples into 9 mL sterile peptone water which resulted in 10^1^ dilutions. Next, 1 mL from 10^1^ dilutions was taken and added with 9 mL sterile peptone water resulting in 10^2^ dilutions. The dilution factors were continued until 10^8^. An amount of 100 μl from each diluted solution was pipetted out and spread evenly on EMB media plates. All media plates were incubated aerobically at 37°C for 24-48 h. From each plate, colonies that appeared as dark centred and flat, with or without metallic sheen were selected and cultured on McConkey and Tryptic Sog Agar (TSA) plates, incubated at 37°C for 24– 48 h. The morphology of the selected colonies was determined by microscopic characterization via bacterial staining.

### Bacterial Identification using Gram Staining

A drop of saline was introduced on the slide containing bacteria inoculates from the colony on the previous EMB media. Then, a bacterial sample was smeared and mixed, followed by heat-fixing by passing the slide through flame on a Bunsen burner quickly. After heat-fixing was done, a few drops of crystal violet solution were flooded onto the slides and allowed to react on the specimen for 1 minute. The slide was rinsed with distilled water and flooded with iodine solution for 1 minute. Excess iodine was rinsed off and followed by adding acetone to decolourize the specimens for 5 seconds. Next, distilled water was used to wash the slide. Safranin was used to counterstain. Excess safranin was washed off with distilled water. The slide was observed under a light microscope (Olympus, USA).

### Antibacterial Susceptibility Testing of Presumptive *E. coli*

Four types of antibiotics comprised of Penicillin, Kanamycin, Chloramphenicol and Tetracycline stocks were prepared. The selection of antibiotic panels was based on the frequent usage in veterinary and human medicine (Ateba et al., 2020). Each of the presumptive *E. coli* isolates was subjected to antibiotic susceptibility using the dilution methods. Bacterial isolates were subjected to the disk diffusion method to determine the zones of inhibition and further confirm their susceptibility to antibiotics (Tendencia, 2004). Multidrug resistance was defined as the nonsusceptibility of an isolate to at least 1 agent in the 4 antibiotics tested (Basak et al., 2016). The isolates were cultured until OD_600_ 0.8-1.0 and 100 *µ*L culture was spread evenly on Mueller Hinton (MH) agar plates. The isolates were tested against four antibiotics as follows: Penicillin (25 *μ*g, 50 *μ*g, 100 *μ*g, 500 *μ*g), Chloramphenicol (25 *μ*g, 50 *μ*g, 100 *μ*g, 500 *μ*g), Tetracycline (25 *μ*g, 50 *μ*g, 100 *μ*g, 500 *μ*g), and Kanamycin (25 *μ*g, 50 *μ*g, 100 *μ*g, 500 *μ*g). Antibiotic disks were immersed in different antibiotic concentrations then aseptically placed using sterile forceps as shown in Table 1. All plates were incubated at 37°C for 24 hours. Halo zones appearance on each plate was observed and recorded.

**Table 1:** Concentration of antibiotics

| Antibiotics | Concentration |  |
| --- | --- | --- |
|  | Recommended Stock<br>Concentration<br>(mg/mL) | Recommended<br>Working Concentration<br>(µg/mL) |
| Penicillin | 100 | 100 |
| Kanamycin | 25 (Dissolve with<br>100% Ethanol) | 25 |
| Tetracycline | 50 | 50 |
| Chloramphenicol | 10 | 10 |

## Results & Discussion

### Isolation and identification of presumptive *E. coli* in culture samples

#### Growth on EMB media

In this study, three types of media comprised of EMB, McConkey and TSA were used to isolate the presumptive *E. coli* from the water and soil samples taken in Kadamaian, Kota Belud. In general, a culture media refers to a solid or liquid preparation that must contain the essential nutrients required by the microorganism used for growth, transport and storing (Bozaslan et al., 2016). Whereas selective media such as EMB and McConkey used in this study refer to a media that allow only certain types of microorganisms to grow and inhibit the growth of other non-targeted microorganisms. Additionally, selective media is not suitable for storage as the specific nutrients contained in the media will become acidic due to the production of secondary metabolites of the microorganisms which then leads to growth inhibition. We used TSA as storage media to keep the isolates viable during the study period.

*Escherichia coli* is a type of bacteria that belongs to the *Enterobacteriaceae* family. They are facultative anaerobic rod-shaped bacteria that do not synthesize oxidase enzyme and at the same time, possessed a fermentative and respiratory. According to Yaratha et al. (2017), *E. coli* are known to ferment lactose to produce hydrogen sulphide which is a weak dibasic acid, however, recent studies show up to 10% of isolates have been periodically identified to be less or non-lactose fermenting.

Eosin methylene blue (EMB) agar provides a rapid and reliable method in identifying *E. coli* from other Gram-negative bacteria (Leininger et al., 2001). According to Leininger et al., (2001), green-metallic sheen colonies will appear on the surface of EMB media which indicates the presence of *E. coli*. The dye contains in the EMB media which are eosin Y and methylene blue act as an inhibitor and also pH indicator suppressed the growth of Gram-positive bacteria and at an acidic pH combine to form a precipitate with green-sheen colors. EMB provides an advantage where it can differentiate *E. coli* from other Gram-negative pathogens quickly and accurately (Leininger et al., 2001). This is because it has properties where it inhibits Gram-positive bacteria and provides a color indication to differentiate between lactose-fermenting species. Darker colonies with a green sheen in EMB agar indicates that the colony is a strong lactose fermenter, while less dark or colorless colonies without green sheen in EMB agar indicates that the colony is a weaker or non-lactose fermenter. The green sheen on the EMB agar plate is very specific to *E. coli*. At an acidic pH, the combination of the eosin and methylene blue dyes form a precipitate which is responsible as the indicator of acid production from the *E. coli* fermenting activities.

According to the results obtained in Figure 1, there was no presence of green sheen. Instead, the EMB agar showed results of dark or black colored colonies only. Generally, this indicated that the colonies were not *E. coli* or it could have been *E. coli* but with weak lactose fermenting abilities. However, the pH of the samples might have played a role in the regulation of the green sheen presence. This statement was supported in a study conducted by Leininger et al. (2001), which used mastitic milk as samples where the outcomes showed that *E. coli* on EMB agar does not necessarily produce green metallic sheen colonies. The green sheen production appears to be sensitive to changes in pH. Therefore, the lack of sheen production could be due to the alkalinity of media interfering with the acidic requirement of EMB agar for the production of the green sheen. Due to the undefined results as shown in Figure 1, another identification method was carried out to grow bacterial colonies on the McConkey agar plates which also acts as a second selective plate for *E. coli* identification.

**Figure 1.**
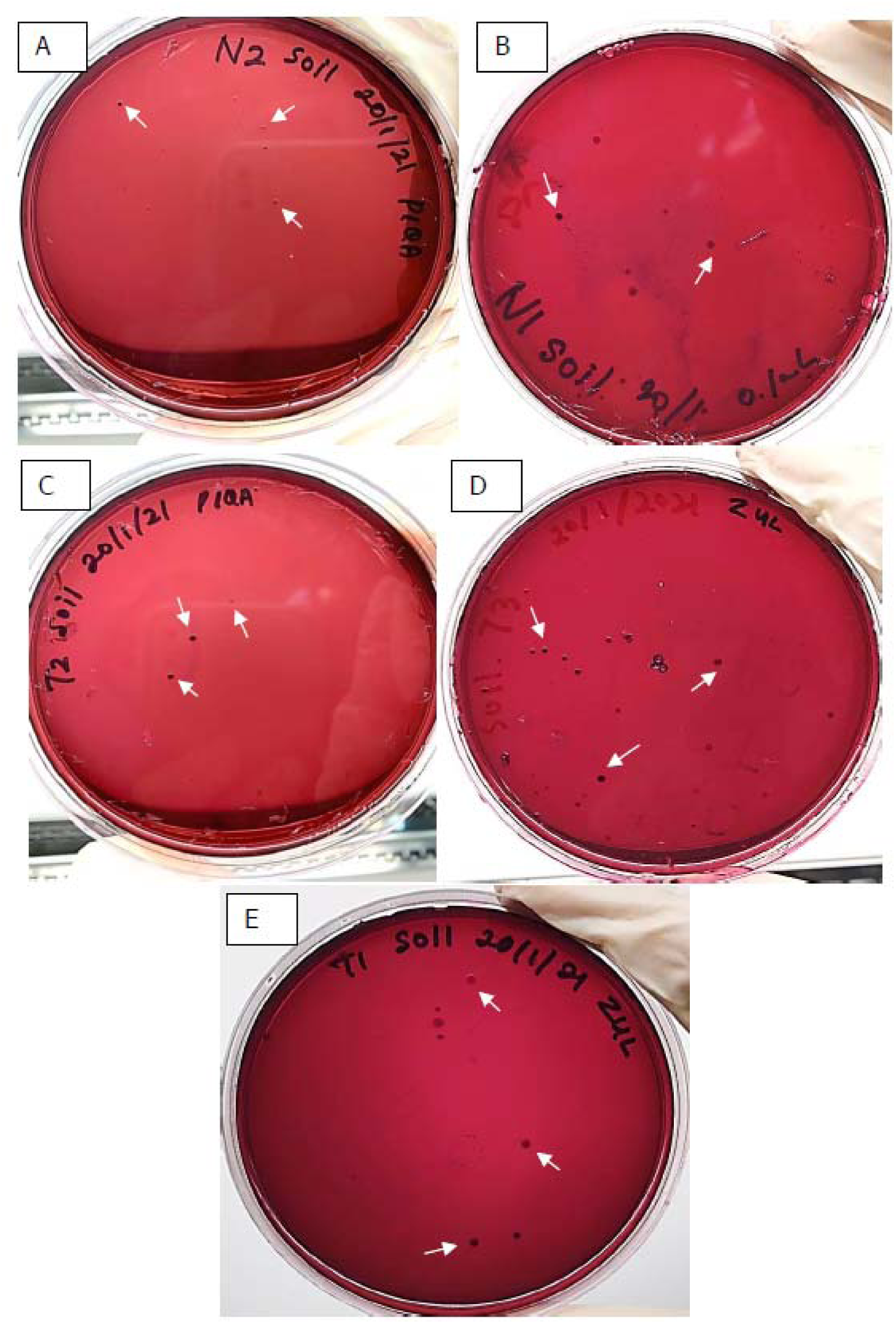
Isolation and identification of presumptive *E. coli* on EMB media. Samples obtained from A & B: Kampung Napungguk (soil sample N1 and N2); C, D & E: Kampung Tinata (soil sample T1, T2, T3). Only soil samples showed brownish colony growth of presumptive *E. coli* bacteria (indicated by white arrows) when incubated on EMB media and there was no growth of *E. coli* on water sample throughout the isolation.

#### *E. coli* growth on McConkey media

Commonly, selective media McConkey is used to distinguish *E. coli* from other Gram-negative bacteria. Same as EMB agar, McConkey also was designed to inhibit the growth of Gram-positive bacteria (Leininger et al., 2001). Lactose fermentation would produce organic acid, lactic acid which then decreases the pH of agar media. Decreased pH indicates an acidic condition. Under an acidic condition, the pH indicator turns into pink color.

Thus, lactose-fermenting-Gram-negatives (lactose-fermenters) will form pink colonies, while non-lactose fermenters will form off-white opaque colonies. Species differences will show different rates of growth even within the lactose fermenters. In short, McConkey can selectively allow only grows Gram-negative bacteria, and those bacteria appeared differently based on their lactose fermenting ability as well as the rate of fermentation and the presence of a capsule or not (Jung B & Hoilat GJ. 2021). Sugumaran et al., (2020) stated that positive growth of lactose-fermenting bacteria will show pink colonies, either with mucoid or without mucoid substances. According to Lupindu (2017), the ability of some *E. coli* strains to ferment different sugars can be beneficial to concentrate the strains of interest. In Figure 2, all isolates from the previous colonies that have been cultured on EMB media showed pink-brownish colonies which specifically indicated the presence of lactose-fermenting bacteria. Further isolation and confirmation were done by inoculating the isolates in TSA media. Generally, TSA media known as Tryptic Soy Agar was used for the isolation and cultivation of various microorganisms either fastidious or non-fastidious types. After the inoculation of isolates from McConkey media was done on TSA media, the cream-colored bacterial growth was observed as shown in Figure 3. The summary of observed characteristics of presumptive *E. coli* was recorded in Table 2.

**Table 2:** Summary of morphological identification and isolation of bacteria using selective media from 3 sampling sites in Kadamaian, Kota Belud.

| Sample | Volume | Dilution | EMB media | MacConkey media | TSA | Presumptive Isolates |
| --- | --- | --- | --- | --- | --- | --- |
| N1 | 0.1ml | 102 | NEG-Dark brownish colonies | POS-Pink colonies | POS-Yellowish Colonies | Enterobacteria spp. |
| N2 | 0.1ml | 102 | NEG-Dark brownish colonies | POS-Pink Colonies | POS-Yellowish Colonies | Enterobacteria spp. |
| T1 | 0.1ml | 102 | NEG-Dark brownish colonies | POS-Light Brown | POS-Yellowish Colonies | Enterobacteria spp. |
| T2 | 0.1ml | 102 | NEG-Dark brownish colonies | POS-Light Brown | POS-Yellowish Colonies | Enterobacteria spp. |
| T3 | 0.1ml | 102 | NEG-Dark brownish colonies | POS-Light Brown | POS-Yellowish Colonies | Enterobacteria spp. |

\*Note POS- labelled for identification of expected *E. coli* growth colony observed on media while NEG- labelled for unmatched characteristics presumes for *E. coli* growth colony on media. N1 – N2 refers to Kg. Napungguk soil samples while T1 – T3 refers to Kg. Tinata soil samples.

**Figure 2.**
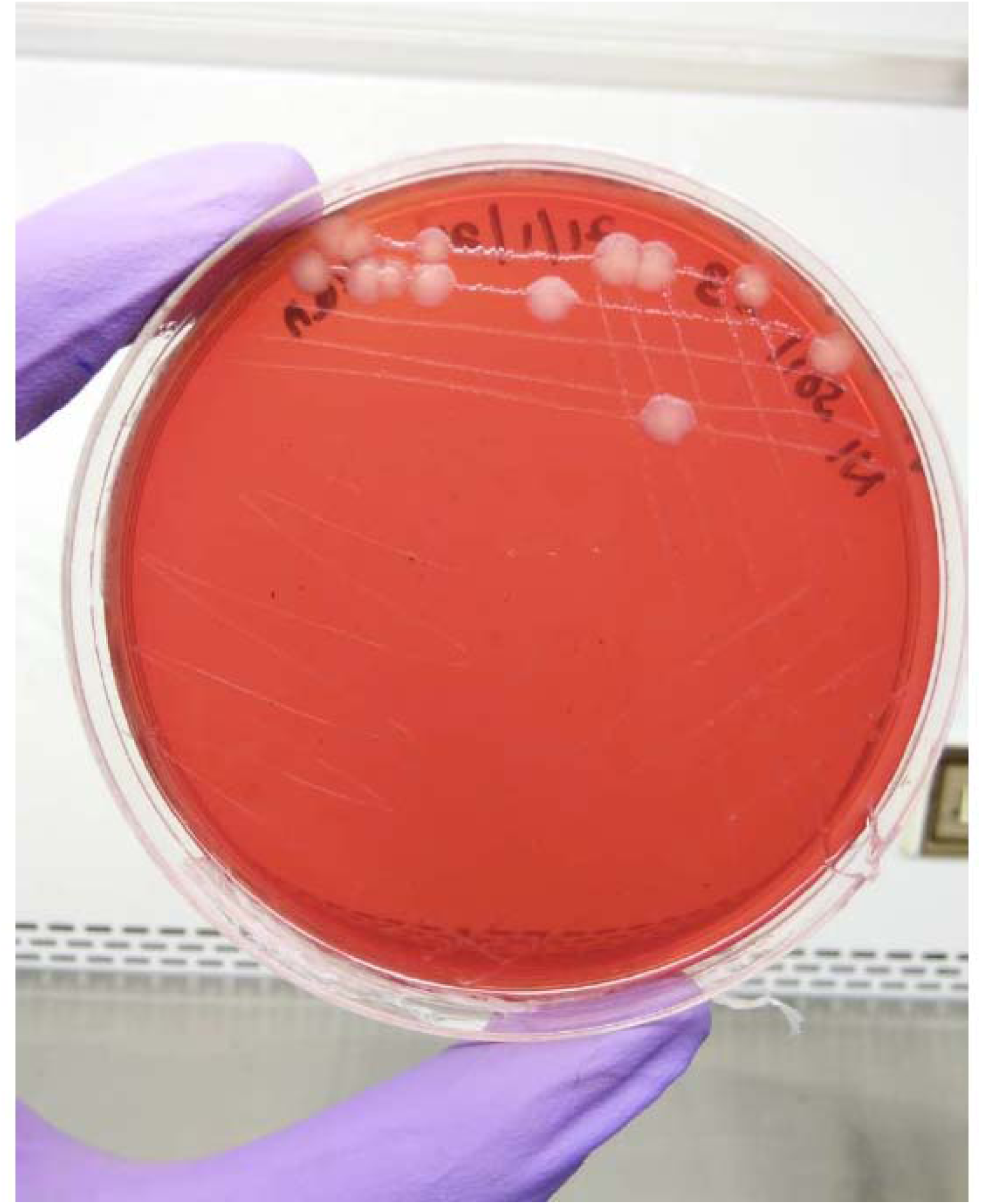
Appearance of pink colonies (indicated by white errors) can be seen on McConkey media for confirmation of presumptive *E. coli* growth on previous EMB plates.

**Figure 3.**
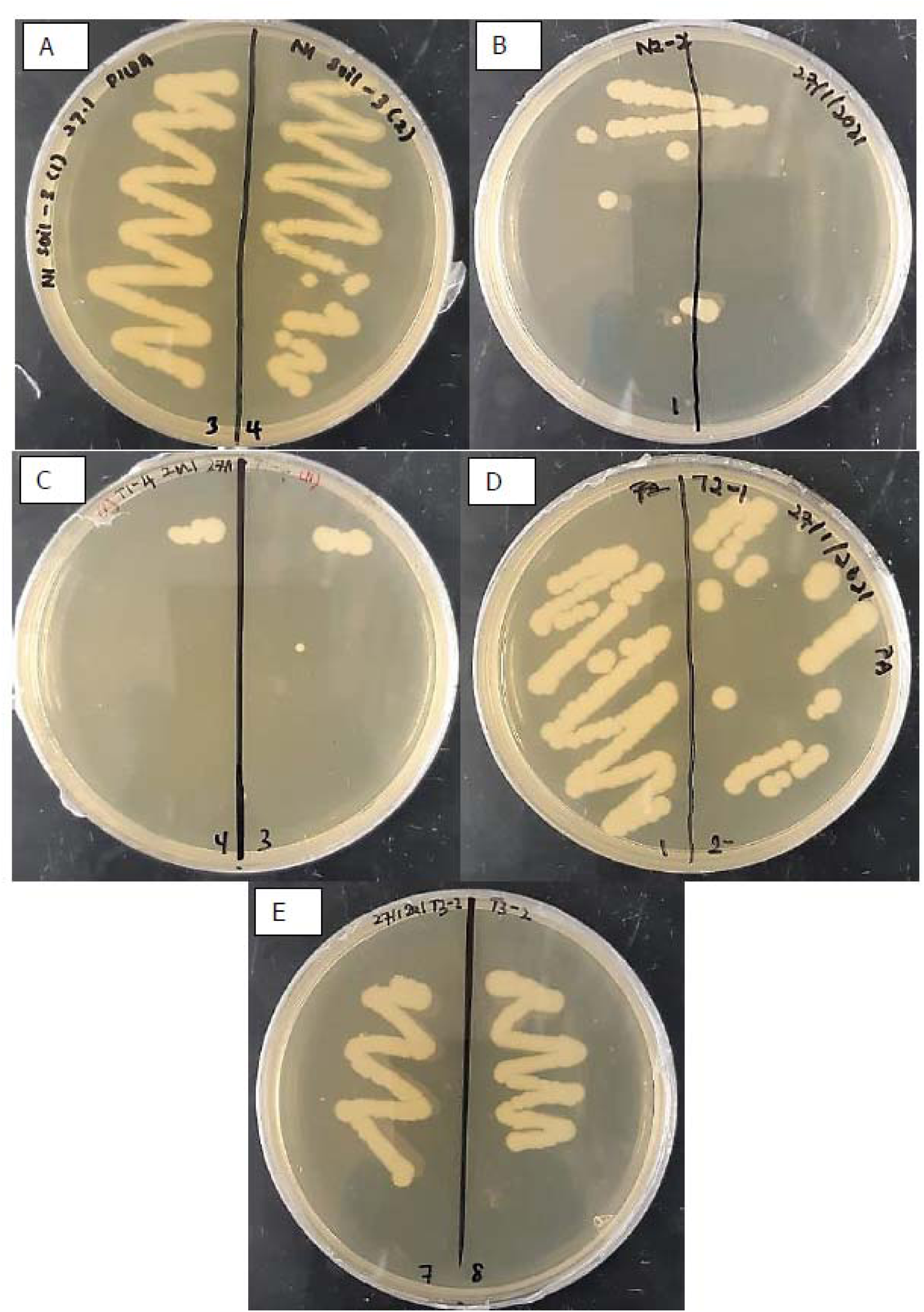
Isolation and identification of presumptive *E. coli* on TSA media. Figure A & B: Kampung Napungguk (soil sample N1 and N2); C, D & E: Kampung Tinata (soil sample T1, T2, T3).

#### Verification of *E. coli* bacteria using the Gram-staining technique

Gram staining is one of the common identification techniques used to distinguish the bacteria according to different components in their cell wall (Bucknner,2016). As Gram-positive bacteria are composed of a thick peptidoglycan layer of about 60 to 90%, therefore after the staining procedure, the bacterial cell will be stained violet or purple. Whereas for Gram-negative bacteria which have a thinner layer of peptidoglycan, the bacterial will appear as red color (Black,2004). Results obtained after the gram staining process showed that all the isolates were stained pink and red color and have a rod-shaped thus confirming that the isolates were presumptive *E. coli* (Figure 4). Summary of the identification and morphological characteristics are shown in Table 3.

**Table 3:** Summary of the identification and morphological characteristics in gram-negative bacteria for each sampling site. ‘X’ refers to the matched characteristics of Gram-staining guides.

| Sampling sites | Replicates | Bacteria | Gram positive | Gram Negative | Morphology |
| --- | --- | --- | --- | --- | --- |
| Kg. Napungguk | N1 | *E. coli* |  | X | Rod-shaped, some aligned 2-3 together, some singular, stained as pink |
|  | N2 | *E. coli* |  | X | Rod-shaped, some aligned 2-3 together, some singular, stained as pink |
| Kg. Tinata | T1 | *E. coli* |  | X | Rod-shaped, some aligned 2-3 together, some singular, stained as pink |
|  | T2 | *E. coli* |  | X | Rod-shaped, some aligned 2-3 together, some singular, stained as pink |
|  | T3 | *E. coli* |  | X | Rod-shaped, some aligned 2-3 together, some singular, stained as pink |

**Figure 4.**
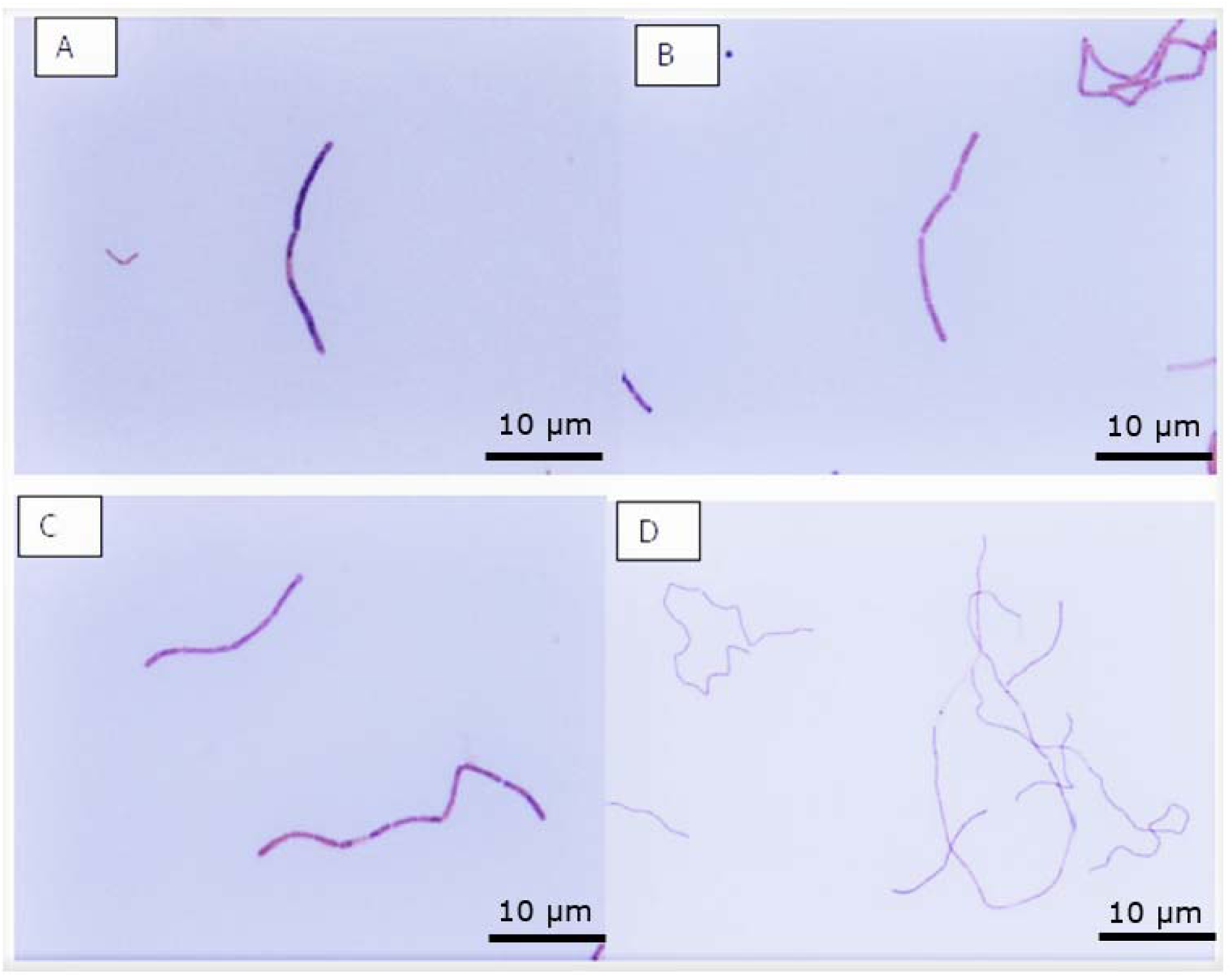
Verification of presumptive *E. coli* bacteria using a gram-staining technique. Images viewed under 1000x magnification using compound light microscope (US Olympus) for *E. coli* gram-negative staining in samples A & B: soil sample from Kg. Napungguk, N1 & N2; C & D: soil sample from Kg. Tinata, T1 & T2.

#### Antibacterial Susceptibility Testing of *E. coli* using antibiotics

Based on Table 4 and Table 5, they represented the antibiotic susceptibility profile of the presumptive *E. coli* isolates. All the isolates were tested against four different antibiotics using disk diffusion to study the susceptibility of the isolates against the subjected antibiotics. Table 5 reveals that the presumptive *E. coli* isolates were most resistant to Chloramphenicol with 91.3%, followed by Penicillin with 82.6%, Tetracycline with 73.9% and Kanamycin with 30.4%. Six out of twenty-three isolates showed resistance to all types of antibiotics in different concentrations. The tested presumptive *E. coli* isolates were most susceptible to Kanamycin with 17.4%, followed by Tetracycline with 13.0% while Penicillin and Chloramphenicol both represented the least susceptible with 4.3%. Analysis of multiple drug resistance of the presumptive *E. coli* reveals that 21 out of 23 isolates exhibit resistance against multiple antibiotics, therefore these isolates can be classified as multidrug resistance. This data showed that almost all of the isolates from five soil samples were obtained from Kg. Tinata and Kg. Napungguk were multidrug resistance which should be alarming news that needs further analysis.

**Table 4:**
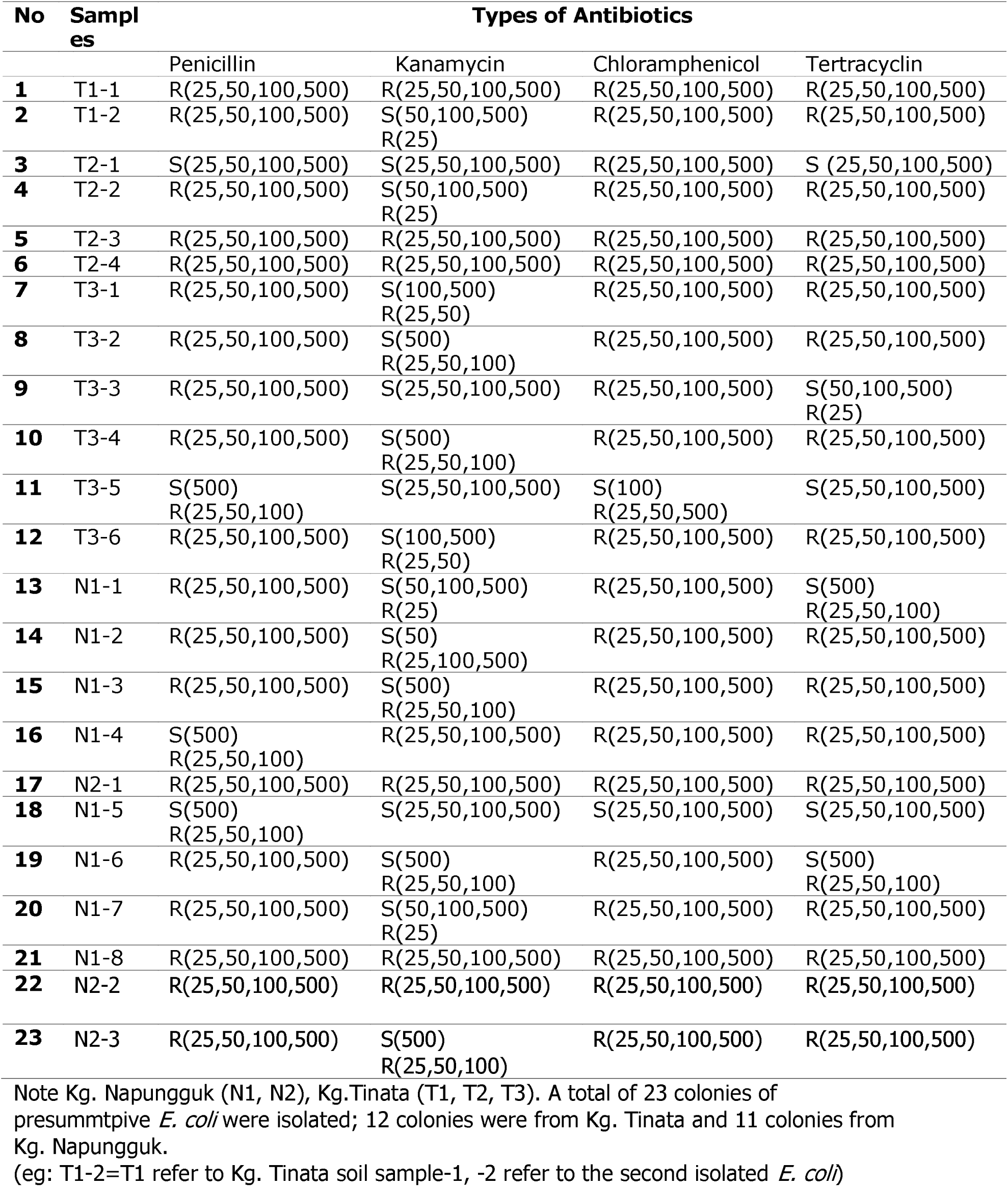
Antibiotics resistance patterns of presumptive *E. coli* isolates from samples Kg. Napungguk (N1-N2) and Kg. Tinata (T1-T3).

| No | Samples | Types of Antibiotics |  |  |  |
| --- | --- | --- | --- | --- | --- |
|  |  | Penicillin | Kanamycin | Chloramphenicol | Tertracyclin |
| **1** | T1-1 | R(25,50,100,500) | R(25,50,100,500) | R(25,50,100,500) | R(25,50,100,500) |
| **2** | T1-2 | R(25,50,100,500) | S(50,100,500)<br>R(25) | R(25,50,100,500) | R(25,50,100,500) |
| **3** | T2-1 | S(25,50,100,500) | S(25,50,100,500) | R(25,50,100,500) | S (25,50,100,500) |
| **4** | T2-2 | R(25,50,100,500) | S(50,100,500)<br>R(25) | R(25,50,100,500) | R(25,50,100,500) |
| **5** | T2-3 | R(25,50,100,500) | R(25,50,100,500) | R(25,50,100,500) | R(25,50,100,500) |
| **6** | T2-4 | R(25,50,100,500) | R(25,50,100,500) | R(25,50,100,500) | R(25,50,100,500) |
| **7** | T3-1 | R(25,50,100,500) | S(100,500)<br>R(25,50) | R(25,50,100,500) | R(25,50,100,500) |
| **8** | T3-2 | R(25,50,100,500) | S(500)<br>R(25,50,100) | R(25,50,100,500) | R(25,50,100,500) |
| **9** | T3-3 | R(25,50,100,500) | S(25,50,100,500) | R(25,50,100,500) | S(50,100,500)<br>R(25) |
| **10** | T3-4 | R(25,50,100,500) | S(500)<br>R(25,50,100) | R(25,50,100,500) | R(25,50,100,500) |
| **11** | T3-5 | S(500)<br>R(25,50,100) | S(25,50,100,500) | S(100)<br>R(25,50,500) | S(25,50,100,500) |
| **12** | T3-6 | R(25,50,100,500) | S(100,500)<br>R(25,50) | R(25,50,100,500) | R(25,50,100,500) |
| **13** | N1-1 | R(25,50,100,500) | S(50,100,500)<br>R(25) | R(25,50,100,500) | S(500)<br>R(25,50,100) |
| **14** | N1-2 | R(25,50,100,500) | S(50)<br>R(25,100,500) | R(25,50,100,500) | R(25,50,100,500) |
| **15** | N1-3 | R(25,50,100,500) | S(500)<br>R(25,50,100) | R(25,50,100,500) | R(25,50,100,500) |
| **16** | N1-4 | S(500)<br>R(25,50,100) | R(25,50,100,500) | R(25,50,100,500) | R(25,50,100,500) |
| **17** | N2-1 | R(25,50,100,500) | R(25,50,100,500) | R(25,50,100,500) | R(25,50,100,500) |
| **18** | N1-5 | S(500)<br>R(25,50,100) | S(25,50,100,500) | S(25,50,100,500) | S(25,50,100,500) |
| **19** | N1-6 | R(25,50,100,500) | S(500)<br>R(25,50,100) | R(25,50,100,500) | S(500)<br>R(25,50,100) |
| **20** | N1-7 | R(25,50,100,500) | S(50,100,500)<br>R(25) | R(25,50,100,500) | R(25,50,100,500) |
| **21** | N1-8 | R(25,50,100,500) | R(25,50,100,500) | R(25,50,100,500) | R(25,50,100,500) |
| **22** | N2-2 | R(25,50,100,500) | R(25,50,100,500) | R(25,50,100,500) | R(25,50,100,500) |
| **23** | N2-3 | R(25,50,100,500) | S(500)<br>R(25,50,100) | R(25,50,100,500) | R(25,50,100,500) |

Note Kg. Napungguk (N1, N2), Kg. Tinata (T1, T2, T3). A total of 23 colonies of presumptive *E. coli* were isolated; 12 colonies were from Kg. Tinata and 11 colonies from Kg. Napungguk. (eg: T1-2=T1 refer to Kg. Tinata soil sample-1, -2 refer to the second isolated *E. coli*)

**Table 5:** Percentage of susceptibility of antibiotic resistance of presumptive *E. coli* isolates from samples Kg. Napungguk (N1-N2) and Kg. Tinata (T1-T3)

| Antibiotic | Disc concentration | Susceptibility |  |  |
| --- | --- | --- | --- | --- |
|  |  | Resistant number (%) | Intermediate number (%) | Sensitive number (%) |
| Penicilin | 25,50,100,500 | 19 out of 23 (82.6%) | 3 out of 23 (13.0%) | 1 out of 23 (4.3%) |
| Kanamycin | 25,50,100,500 | 7 out of 23 (30.43%) | 12 out of 23 (52.2%) | 4 out of 23 (17.4%) |
| Chloramphenicol | 25,50,100,500 | 21 out of 23 (91.3%) | 1 out of 23 (4.3%) | 1 out of 23 (4.3%) |
| Tetracycline | 25,50,100,500 | 17 out of 23 (73.9%) | 3 out of 23 (13.0%) | 3 out of 23 (13.0%) |

The emergence of *E. coli* that acquired multidrug-resistant (MDR) had caused antibiotic management problems in healthcare centres and hospitals in Malaysia. According to Odonkor & Addo (2018), infectious disease control is under pressure by the rise in the number of microorganisms that are resistant to antimicrobial agents. Due to the failure of the disease responding to the subjected conventional treatment, thus resulting in prolonged illness and greater risk of death. The main cause of the emergence of MDR is due to genetic mutation in bacteria (Laxminarayan, 2003). Odonkor & Addo, (2018), stated that the greater the duration of exposure of the antibiotic on the bacteria, the greater the risk of the development resistance towards antibiotics.

Based on the result, the frequency of Chloramphenicol resistance was the highest among the isolates as compared to the Penicillin, Kanamycin and Tetracycline. According to Fernandez (2012), historically, Chloramphenicol is a wide range of antibiotics purposely to treat all major food-producing animals and is currently used in humans and pets.

Chloramphenicol is the D-threo isomer of a molecule in which consisting of a p-nitrobenzene ring connected to a dichloroacetyl tail via a 2-amino-1,3-propanediol moiety (Dinos et al., 2016). The mechanisms of action Chloramphenicol cause the bacteriostatic activity by blocking the bacterial protein synthesis (Dinos et al., 2016). Furthermore, Chloramphenicol also exhibits bactericidal activity in which against the three common causes of meningitis such as *Haemophilus influenza, Streptococcus pneumoniae* and *Neisseria meningitis* (Dinos et al., 2016). A study done by Peterson (2004) on the development of antibiotic resistance in integrated fish farms in Asia revealed that the antimicrobial agent residue and animal manure retrieved from the farm and was shed directly into the fish pond as fertilizer has contributed to the formation of antibiotic resistance. A result from the research conducted by Geidam, (2012) also revealed the presence of multidrug resistance in chicken due to the use of antibiotics in chicken food.

In our case, the emergence of Chloramphenicol resistance is possibly due to agricultural activities especially in-related to animal husbandry around the Kadamaian area. Data on the distribution of Sabah ruminant breeders in 2016 from the Livestock Census showed that Kota Belud has the highest number of ruminant breeders with 391 in total (Data.gov.my, 2018). Due to this, the soil and water sources around Kadamaian might have been polluted by the cow’s dung especially if there was no specific area for farming. However, a detailed study needs to be done to identify the type of *E. coli* isolated around Kedamaian areas using sequencing analysis.

The second most resistant antibiotic is Penicillin with a resistance number of 82.6%. This finding is in agreement with the study conducted by Kazemnia et al., (2014) which indicates that Penicillin is one of the most commonly used antibiotics besides Erythromycin, Nalidixic acid, Cephalexin, Amoxicillin, Ampicillin and Ciprofloxacin that tend to exhibit antibiotic-resistance. According to Bouza & Cercenado, (2002); Nordmann P., (1998), *Enterobacteriaceae* especially *E. coli* is among the group of bacteria with high rates of penicillin resistance.

Our results showed that Tetracycline with 73.9% resistance numbers possessed the third-highest among the other subjected antibiotics. According to Bryskier A., (2005), for more than half of the century, Tetracycline has been used to treat infectious diseases and also as a growth-promoting agent in food animal production systems (Dupont HL & JH., 1987; Schnappinger & Hillen, 1996). Landers et al. (2012) stated that Tetracycline is widely used in animal husbandry. Tetracycline exhibit a bactericidal mechanism by disrupting the protein synthesis when it binds to the 30S ribosomal subunit which blocks aminoacyl-t-RNA binding to the ribosomal A thus causing the polypeptides to not synthesized (Adesoji et al., 2015). Kota Belud has the highest ruminant breeders, thus the emergence of Tetracycline-resistance could be related to agricultural activities especially livestock farming in the residential area. Excessive livestock waste as a result of an intensive farming system could lead to soil and water pollution in the Kadamaian area.

Table 5 showed that Kanamycin represents the lowest resistance numbers among the other antibiotics and this is might due to the uncommon use of this type of antibiotic in clinical practice or veterinary medicine. According to Chaudhry S.B. (2016), Amikacin and Kanamycin are mainly used for treating Tuberculosis disease also known as multi-drug resistant tuberculosis (MDR-TB) in developing countries. Brossier et al.,(2017) stated that the resistance to second-line injectable drugs (SLID) such as Capreomycin and Kanamycin is due to the mutation that occurred in rrs region 1400, tlyA and eis promoter.

## Conclusion

The overall result revealed that 21 out of 23 (91.2%) of the isolates showed resistance to at least three subjected antibiotics thus making them a serious health threat. This also indicates that 91.2% of the isolates are multiple antibiotics resistant. In this study, there are several limitations in identifying the isolates as an observation on phenotypic characteristics alone is not enough to confirm the presence of the bacteria. Thus, further investigation and identification using biochemical tests and sequencing analysis on that particular isolates should be carried out. The finding of this study is very important as the sites were residential areas and also tourist locations where many people come to visit Kadamaian, Kota Belud for leisure activities. This finding can be used by the health department to analyse the soil and water samples from time to time to study the emergence of multidrug pathogens that could spread in the area. In conclusion, further identification and confirmation of bacterial strains via molecular methods by the identification of 16S rRNA is highly recommended to study the emergence of multidrug pathogens that could spread in the area.

## Data Availability

All data produced in the present work are contained in the manuscript

## Acknowledgement

Gratitude to Biotechnology Research Institute (BRI) for allowing us to use the resources and facilities for this experiment. We would like to thank Miss Nurfaezah Adam for helping us with the collection of soil and water samples.

